# The impacts of United States foreign development assistance reductions on health system building blocks at healthcare facilities in Zambia: a mixed-methods study

**DOI:** 10.1101/2025.10.06.25337414

**Authors:** Lucy K. Tantum, Darcy M. Anderson, Hikabasa Halwiindi, Michael Herce, Michael T. Mbizvo, Miles A. Kirby, Ryan Cronk

## Abstract

The rapid reduction in foreign development assistance from the United States and other countries has led to disruptions in essential global health programming. Countries that previously received development assistance, such as Zambia, may experience weakening of health system capacity due to program cancellations. This study aimed to describe the impacts of USAID program defunding on healthcare delivery in Zambia. We conducted a cross-sectional survey at 34 healthcare facilities in three districts of Zambia in April and May 2025. Through facility-level assessments and individual surveys with 330 healthcare workers, we identified changes in health system building blocks that may have arisen from program defunding. The facility-level assessment found that 71% of healthcare facilities (n=24) experienced changes related to funding cuts within the previous three to four months. In open-ended surveys, healthcare workers reported impacts on the health system, including stock-outs of essential medicines, diagnostic tests, and infection control supplies; layoffs of US-supported healthcare workers; and reduced ability to work with electronic medical records. Workers described how these changes impacted workplace morale, patient satisfaction, and their ability to deliver essential services. This study reveals the immediate consequences of defunding foreign assistance, which, if left unaddressed, may weaken health systems and worsen health outcomes. To mitigate these impacts, country governments and partner organizations should prioritize interventions and investments that strengthen health systems, such as expanding healthcare revenue streams and building workforce capacity. In the wake of funding cuts, health system strengthening can reduce countries’ reliance on foreign assistance and improve population health.

## Introduction

United States official development assistance has been an important source of economic support for global health, comprising 29% of all health development assistance in low- and middle-income countries in 2023 (1). US development assistance has supported a wide range of global health initiatives, including programs addressing maternal and child health, HIV/AIDS, malaria, tuberculosis, and health system strengthening (2). In January 2025, the US federal government announced a pause in foreign development assistance and began suspending or eliminating United States Agency for International Development (USAID) programs (3). Subsequently, in March 2025, the administration announced the cancellation of 83% of USAID programs and the intent to dissolve the Agency (4,5). A small proportion of these programs was later reinstated through the US State Department (6,7). Following the US funding cuts, several European countries also reduced their foreign assistance programs, resulting in an overall estimated 22% drop in development assistance for health from 2024 to 2025 (8,9).

This reduction in foreign assistance funding has had widespread implications for health systems globally (10,11). USAID-supported organizations have described immediate disruptions to health-related programming, including worker layoffs, stockouts of essential medicines and medical supplies, and reduced ability to deliver services such as immunization and diagnostic testing (8,12,13). In the long term, if gaps in funding are not addressed, foreign assistance cuts will lead to gaps in service delivery that could increase global mortality and reverse progress toward universal health coverage (14–16). A forecasting analysis estimated that reductions in USAID funding could lead to over 600,0000 deaths in children under age 5 and over 1.7 million deaths in all ages by 2030, drawing from estimates of USAID’s health impacts across all program areas (17). Other models have projected that reductions in foreign assistance could result in over 50,000 excess child deaths from malaria, over 126,000 excess child deaths from diarrhea, and over 62,000 excess deaths from tuberculosis in one year (18).

Indirect impacts of reductions in foreign assistance may also arise from changes at the healthcare facility level. Through its programming for global development and maternal-child health, USAID has supported infectious disease testing and treatment, health workforce development, and healthcare facility infrastructure improvement, including programs advancing water, sanitation and hygiene (WASH) (19,20). Reduced WASH infrastructure provision in healthcare facilities may increase the risk of healthcare-associated infections, prolonging illness and driving up healthcare costs (21,22). Impacts on the quality and timeliness of healthcare may also arise from shortages of essential medicines and healthcare worker layoffs. Studies of healthcare utilization during epidemics indicate that substandard conditions at facilities may also lead patients to delay or avoid seeking healthcare (23,24).

The effects of defunding will disproportionately impact health systems in countries that were previously receiving large amounts of USAID funding, including Zambia (9). In the 2023 fiscal year, USAID allocated $360 million to programs in Zambia, including $188 million for the HIV/AIDS sector, $66 million for the basic health sector (including tuberculosis, malaria, and nutrition), and $20 million for maternal-child health and family planning (25). In comparison, at the time of this publication, USAID had allocated $135 million to programs in Zambia in the 2025 fiscal year, and recissions may contribute to further cuts (4,25). As foreign development assistance comprises a large portion of its national health budget, overall health spending in Zambia is projected to decline by 10-15% in 2025 compared to 2022 (9). Funding reductions may impede health system strengthening in Zambia. Despite efforts to recruit and train healthcare workers, Zambia still faces a severe healthcare worker shortage, with an estimated 1:12,000 doctor-to-patient ratio (compared to an ideal ratio of 1:5000), and 1:14,900 nurse-to-patient ratio (compared to an ideal ratio of 1:700) (26). Many healthcare facilities lack essential infrastructure: a 2024 UNICEF report estimated that, among non-hospital facilities in Zambia, 68% had basic water services, 1% had basic sanitation services, and 13% had basic waste management services in 2023 (27).

To date, few studies have collected and reported primary data on the health system impacts of foreign assistance funding cuts in Zambia or other low- and middle-income settings. Research on this topic can inform strategies to maintain continuity in healthcare delivery despite variable funding. The objectives of this study were to describe how reductions in foreign development assistance have impacted the health system in Zambia, and assess the implications of these changes for the health and well-being of patients and workers in healthcare facilities.

## Methods

### Study design

A team of American and Zambian researchers surveyed staff at healthcare facilities in Zambia and performed a cross-sectional assessment of health service delivery to evaluate changes experienced in the time period shortly after USAID program suspensions began. Data collection took place in April and May 2025, three to four months after the US government announced the initial USAID funding freeze and program review (3). This study was part of a broader evaluation of WASH and healthcare worker well-being at healthcare facilities in Zambia. With facility-level data, we characterized the general impacts of defunding on facility operations and described the availability of WASH services, essential medicines, and medical equipment. We analyzed individual healthcare workers’ open-ended survey responses to assess changes in health system inputs. Using the World Health Organization (WHO) Building Blocks of Health Systems framework, we classified changes reported in healthcare worker surveys that related to one or more building blocks: service delivery, health workforce, access to essential medicines, health information systems, financing, and governance (28).

### Study sites and population

Study sites were government-run healthcare facilities in Luwingu District and Lupososhi District, Northern Province, and Lusaka District, Lusaka Province, Zambia. These districts were purposively selected as part of a program evaluation led by the non-governmental organization World Vision. We selected districts that had previously received the World Vision program or could serve as a non-program comparison group. Within each district, we purposively selected study sites based on urban/rural status and facility type to obtain data from a range of healthcare contexts. Eligible facility types included health posts, health centers, mini hospitals (which offer diagnostic services, minor surgical services, and inpatient care), and first-level hospitals (which offer medical and surgical services and receive referrals from mini hospitals and health centers) (29).

At each facility, we purposively selected one knowledgeable staff member, such as a medical director or superintendent, nurse in-charge, or environmental health technician, to participate in an assessment of infrastructure and service provision. When needed, we consulted additional staff for information that the original respondent was unable to provide. We selected additional healthcare workers to participate in individual-level surveys. All individuals employed by the healthcare facility, including clinical and non-clinical staff, were eligible to participate in surveys. Many of the rural health posts and clinics in our sample had fewer than 10 staff members present on the day of data collection; at these facilities, we selected all available staff to participate in the survey. At larger healthcare facilities with more staff present, we used a purposive and convenience sampling approach to select individuals based on their staff role, clinical department, and availability to participate. The research team visited each department and invited several workers of different roles to participate in the survey. At these larger facilities, the target sample size was 15-20 participants.

### Data collection

At each selected healthcare facility, an American researcher assessed service delivery by speaking with a knowledgeable staff member and directly observing WASH infrastructure, including water systems, hand hygiene stations, toilets, and waste management facilities. The facility-level assessment contained questions on types of healthcare services provided and typical patient volumes; availability and functionality of WASH infrastructure; and availability of essential medicines and equipment. We determined availability of seven essential medicines (ciprofloxacin, amoxycillin, paracetamol, oral rehydration salts, any first-line tuberculosis treatment, any first-line antimalarial, and any antiretroviral) and six medical equipment (weighing scale, thermometer, stethoscope, blood pressure cuff, refrigerator, and computer with internet access), drawing from WHO guidelines for essential medicine and service delivery indicators (28).

The facility-level assessment also asked a question about US foreign development assistance reductions: ‘Has the facility experienced changes due to recent USAID funding cuts?’ If respondents answered affirmatively, researchers asked an open-ended question about the type of impact experienced and coded the best-fitting response category on the survey form. The response categories were developed during a survey pre-test at three healthcare facilities. During pre-testing, we collected preliminary information about types of impacts experienced at facilities through open-ended survey questions and informal discussions with staff. Researchers could also select an ‘other’ category and write a text description for impacts that did not fit into any categories. Researchers verbally administered all assessment questions and recorded responses in a Kobo Toolbox form on a tablet.

We collected additional data on changes in healthcare facilities through a survey of individual healthcare workers. Survey questions were part of a larger assessment of WASH-related well-being among healthcare workers. Zambian researchers verbally administered an initial, open-ended question about whether the facility had experienced any changes since the start of the foreign development assistance freeze in January 2025. Depending on the participant’s response, researchers then asked additional open-ended probing questions. Researchers were given a list of probing questions and independently decided which, if any, to ask the participant. These optional probes asked if the availability of essential medicines or WASH services had changed; whether any changes had occurred because of USAID defunding; and how changes had impacted worker and patient well-being. However, the survey form did not direct researchers to ask about every health system building block component. Participants responded verbally, and researchers transcribed or paraphrased responses in text using a Kobo Toolbox form.

### Data analysis

With data from healthcare facility assessments, we generated descriptive statistics for facility characteristics, such as facility type, size, and availability of essential medicines and equipment. We aggregated data on WASH infrastructure and supply availability to determine the proportion of facilities with a basic level of each WASH service, in accordance with criteria from the WHO/UNICEF Joint Monitoring Program (27). This framework classifies each WASH service as ‘basic,’ ‘limited,’ or ‘no service’ based on factors such as infrastructure type, location, and functionality (27). Facility-level assessments also asked a knowledgeable staff member to reflect on impacts that the facility had experienced from USAID defunding. We used these assessment responses to characterize types of impacts reported and whether these impacts affected service delivery. When characterizing the types of impacts reported in the facility-level assessment, we re-categorized impact types recorded as ‘other’ by placing them into existing categories or creating new categories if needed. We used R software (Version 4.4.2) for statistical analysis.

To assess elements of the health system that may be impacted by USAID defunding, we performed thematic analysis of open-ended questions from surveys of individual healthcare workers. To classify changes reported by healthcare workers, we created a codebook with codes and definitions adapted from the WHO Building Blocks of Health Systems framework (28). We classified each participant-reported change based on the health system building block to which it most closely corresponded. We created additional codes to categorize downstream impacts on patient and healthcare worker health and well-being, and to classify responses of no change or a change unrelated to the health system. We coded survey responses using NVivo software (Version 14). Within each code, we reviewed and synthesized responses to describe how defunding of foreign development assistance may affect health system building blocks and patient/worker well-being at Zambian healthcare facilities.

### Ethics

At all healthcare facilities, we obtained permission from the medical director or other authority before initiating data collection. Healthcare workers provided written informed consent for participation in surveys. The study protocol was reviewed by the University of North Carolina IRB and was determined to be exempt from further review (reference no. 24-0733). The University of Zambia Humanities and Social Sciences Research Ethics Committee (HSSREC) IRB reviewed and approved the protocol (reference no. 00006464). The protocol also received clearance from Zambia’s National Health Research Authority, Ministry of Health, and District Health Offices for Luwingu, Lupososhi, and Lusaka.

## Results

### Facility-level characteristics and funding cut impacts

We assessed infrastructure and service delivery at 34 healthcare facilities across three districts of Zambia (Table 1). Facilities were health posts, health centers, mini hospitals, and first-level hospitals. Sixty-two percent of facilities (n=21) were in rural areas, while 38% (n=13) were urban. Most facilities (59%, n=20) provided both inpatient and outpatient services, with others providing only outpatient services. Most facilities had high volumes of outpatient visits, typically screening a median of 590 outpatients per month. Those that provided inpatient services typically admitted a median of 22 inpatients per month. In our evaluation of essential medicines and medical equipment availability, 29% percent of facilities (n=10) had all medicines available and 53% (n=18) had all equipment available on the day of the assessment. On average, facilities had six of seven medicines available and five of six equipment available. Most facilities (62%, n=21) used electricity from the national grid as their main energy source, while others primarily or solely used solar electricity. Facilities reported that they usually had electricity for between five and 24 hours each day, with a mean of 18 hours.

**Table 1.** Administrative characteristics and essential equipment and medicine availability at healthcare facilities in Zambia, 2025 (n=34).

| Healthcare facility characteristics | Number<br>(percent) |
| --- | --- |
| *Basic characteristics* |  |
| Facility type: |  |
| Health center | 14 (41.2%) |
| Health post | 12 (35.3%) |
| First-level hospital | 5 (14.7%) |
| Mini hospital | 3 (8.8%) |
| Administrative district: |  |
| Luwingu District | 13 (38.2%) |
| Lusaka District | 12 (35.3%) |
| Lupososhi District | 9 (26.5%) |
| Location type: |  |
| Rural | 21 (61.8%) |
| Urban | 13 (38.2%) |
| Services provided: |  |
| Both inpatient and outpatient services | 20 (58.8%) |
| Outpatient services only | 14 (41.2%) |
| Typical number of inpatients admitted per month, median [Min, Max] | 22 [5, 250] |
| Typical number of outpatients seen per month, median [Min, Max] | 590 [60, 18000] |
| *Essential equipment, medicines, and infrastructure* |  |
| 6 out of 6 essential equipment* are available and functional | 18 (52.9%) |
| 7 out of 7 essential medicines** are available | 10 (29.4%) |
| Primary energy source |  |
| Grid electricity | 21 (61.8%) |
| Solar electricity | 13 (38.2%) |

\* Essential equipment assessed: Weighing scale, thermometer, stethoscope, blood pressure cuff, refrigerator, and computer with internet access.
\*\* Essential medicines assessed: Ciprofloxacin, amoxycillin, paracetamol, oral rehydration salts, any first-line tuberculosis treatment, any first-line antimalarial, and any antiretroviral.

We assessed WASH service levels according to criteria from the WHO/UNICEF Joint Monitoring Programme (27) (Table 2). Ninety-four percent of facilities (n=32) had a basic water service, defined as an improved water source such as a piped water system, standpipe, or borehole, on facility premises. However, many facilities did not have continuous water service, with 41% (n=14) reporting that water was available from their main source for fewer than 24 hours each day. Fifteen percent of facilities (n=5) had a basic sanitation service; these facilities had toilets or ventilated improved pit latrines with at least one dedicated staff toilet, at least one sex-separated toilet, and toilets accessible for individuals with limited mobility. A basic hand hygiene service, defined as functional hand hygiene facilities at all points of care and toilets, was available at 15% of facilities (n=5). A basic waste management service, including safe segregation and disposal of sharps and infectious waste, was present at 26% of facilities (n=9). Forty-one percent of facilities (n=14) had a basic environmental cleaning service, defined as having cleaning protocols and training of all cleaning staff. One facility (3%) had a basic level of all five WASH services.

**Table 2.**
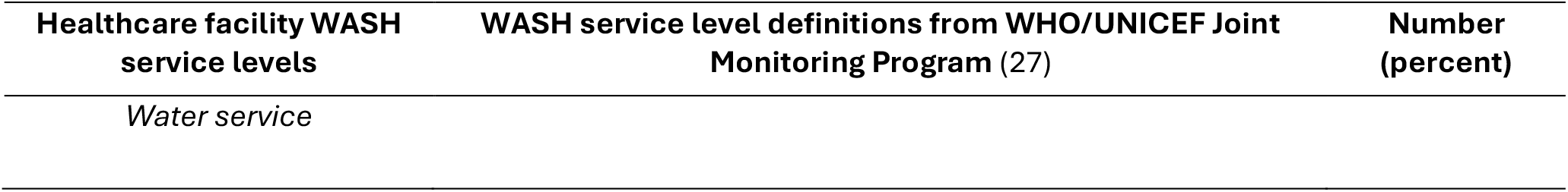
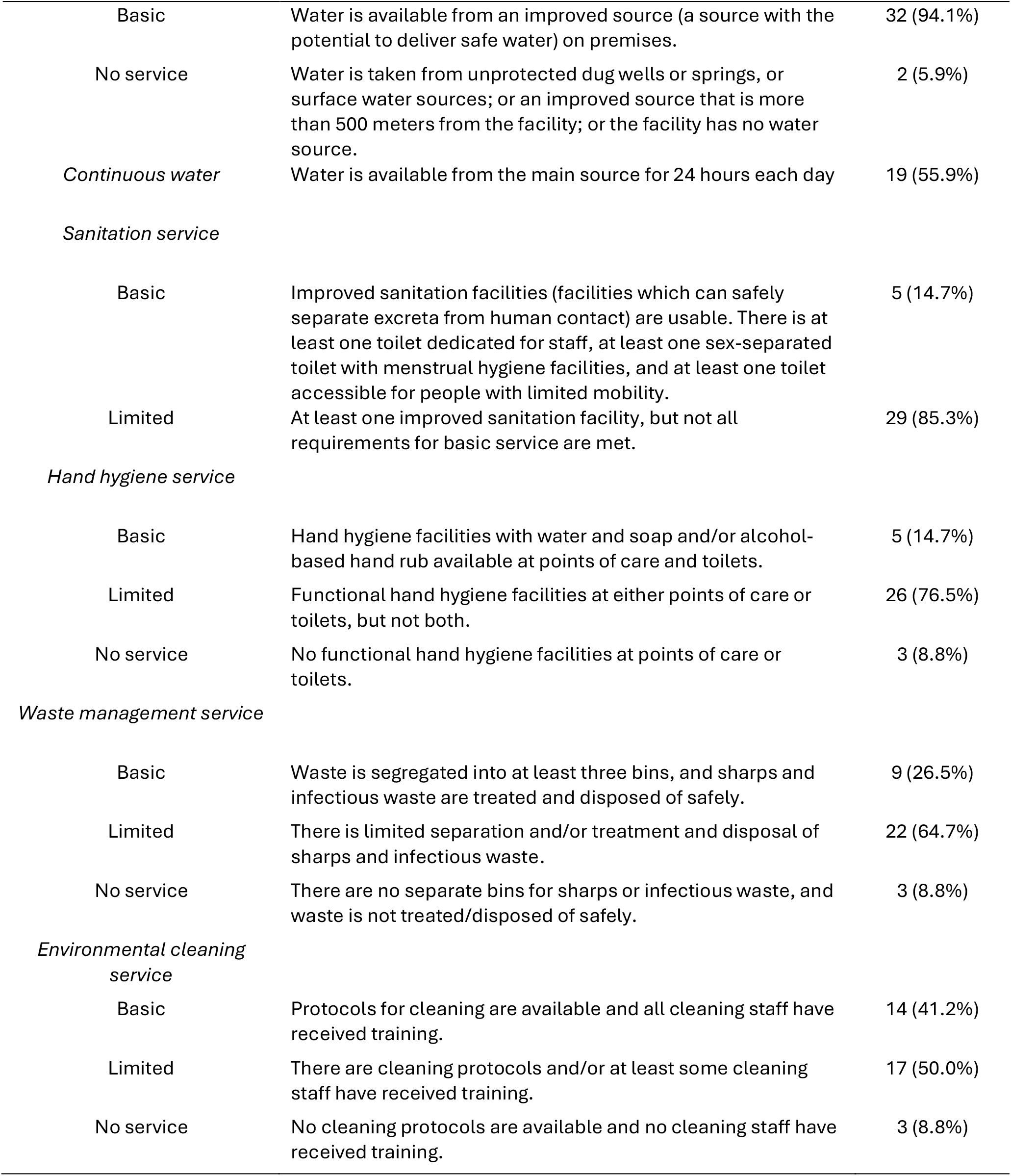
WASH (Water, sanitation, hygiene, cleaning, and waste management) service levels at healthcare facilities in Zambia, 2025 (n=34). WASH service levels are defined according to WHO/UNICEF Joint Monitoring Program criteria (27).

| Healthcare facility WASH service levels | WASH service level definitions from WHO/UNICEF Joint Monitoring Program (27) | Number (percent) |
| --- | --- | --- |
| *Water service* |  |  |
| Basic | Water is available from an improved source (a source with the potential to deliver safe water) on premises. | 32 (94.1%) |
| No service | Water is taken from unprotected dug wells or springs, or surface water sources; or an improved source that is more than 500 meters from the facility; or the facility has no water source. | 2 (5.9%) |
| *Continuous water* | Water is available from the main source for 24 hours each day | 19 (55.9%) |
| *Sanitation service* |  |  |
| Basic | Improved sanitation facilities (facilities which can safely separate excreta from human contact) are usable. There is at least one toilet dedicated for staff, at least one sex-separated toilet with menstrual hygiene facilities, and at least one toilet accessible for people with limited mobility. | 5 (14.7%) |
| Limited | At least one improved sanitation facility, but not all requirements for basic service are met. | 29 (85.3%) |
| *Hand hygiene service* |  |  |
| Basic | Hand hygiene facilities with water and soap and/or alcohol-based hand rub available at points of care and toilets. | 5 (14.7%) |
| Limited | Functional hand hygiene facilities at either points of care or toilets, but not both. | 26 (76.5%) |
| No service | No functional hand hygiene facilities at points of care or toilets. | 3 (8.8%) |
| *Waste management service* |  |  |
| Basic | Waste is segregated into at least three bins, and sharps and infectious waste are treated and disposed of safely. | 9 (26.5%) |
| Limited | There is limited separation and/or treatment and disposal of sharps and infectious waste. | 22 (64.7%) |
| No service | There are no separate bins for sharps or infectious waste, and waste is not treated/disposed of safely. | 3 (8.8%) |
| *Environmental cleaning service* |  |  |
| Basic | Protocols for cleaning are available and all cleaning staff have received training. | 14 (41.2%) |
| Limited | There are cleaning protocols and/or at least some cleaning staff have received training. | 17 (50.0%) |
| No service | No cleaning protocols are available and no cleaning staff have received training. | 3 (8.8%) |

In the facility-level assessment, 71% of respondents (n=24) reported that their facility had experienced changes related to recent USAID funding cuts (Table 3). Among facilities that had experienced changes, the most common reported impact was on staffing levels, with 42% (n=10) of respondents reporting that the number of non-medical staff at the facility had changed and 33% (n=8) reporting that the number of medical staff had changed. Medical staff were defined as staff who provided direct care to patients (e.g., nurses, doctors, counselors) while non-medical staff were defined as staff who were employed by the facility but did not provide direct care (e.g., cleaners, laboratory staff, environmental health technicians). Other respondents described changes in the availability of essential medicines, supplies, and WASH services; patient health-seeking behavior and attendance at the facility; facility budgets; and transport of laboratory samples. Eighty-three percent of respondents who reported changes (n=20) said that these changes had affected the facility’s ability to deliver healthcare services.

**Table 3.**
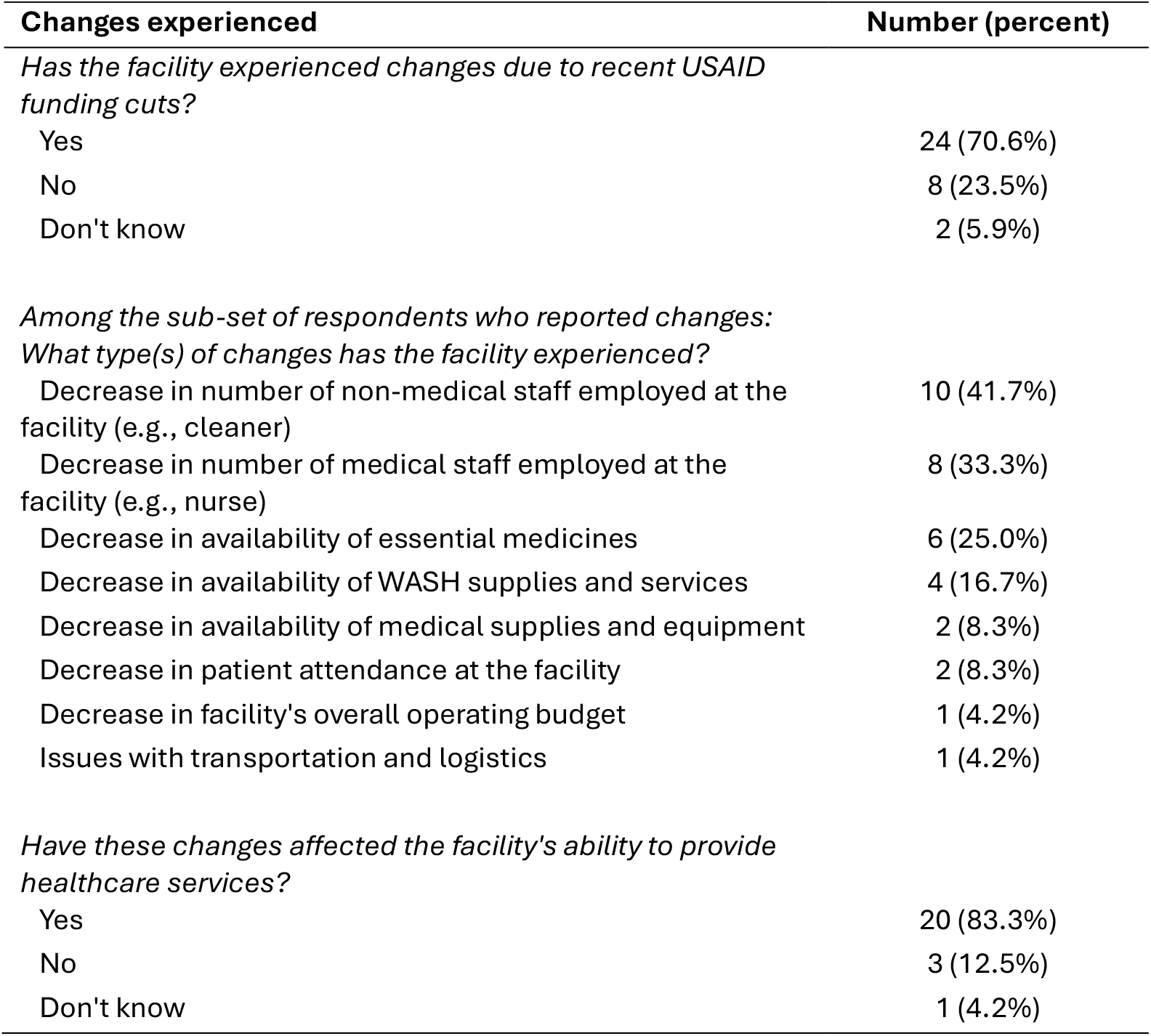
Changes reported at healthcare facilities in Zambia (n=34) related to USAID defunding, 2025.

### Health system impacts observed by healthcare workers

Healthcare workers (n=330) participated in individual-level surveys about changes that had occurred at healthcare facilities from January 2025 through the day of data collection in April or May 2025. Survey participants represented a range of medical and non-medical staff roles (Table 4). Approximately half of participants (n=158, 48%) reported that no changes had occurred or only described changes unrelated to health systems. Among the 172 participants (52%) who described changes in one or more health system building blocks, many described detrimental impacts that may have occurred in connection with USAID funding cuts. Some also described implications of these system-level changes for patient and worker well-being. Figure 1 depicts a conceptual model for changes in health system inputs, healthcare worker-reported immediate impacts of these changes, and potential long-term impacts.

**Table 4.** Characteristics of participants (n=330) in healthcare facility staff survey, Zambia, 2025.

| Participant characteristics | Number (percent) |
| --- | --- |
| Role: |  |
| Doctor, medical licentiate, or clinical officer | 32 (9.7%) |
| Nurse | 100 (30.3%) |
| Other medical staff | 69 (20.9%) |
| Counselor or community health assistant | 36 (10.9%) |
| Cleaner, maid, or general worker | 56 (17.0%) |
| Other non-medical staff | 37 (11.2%) |
| Gender: |  |
| Female | 218 (66.1%) |
| Male | 112 (33.9%) |
| Healthcare facility type: |  |
| Health center | 148 (44.8%) |
| First-level hospital | 113 (34.4%) |
| Health post | 37 (11.2%) |
| Mini hospital | 32 (9.7%) |

**Figure 1.**
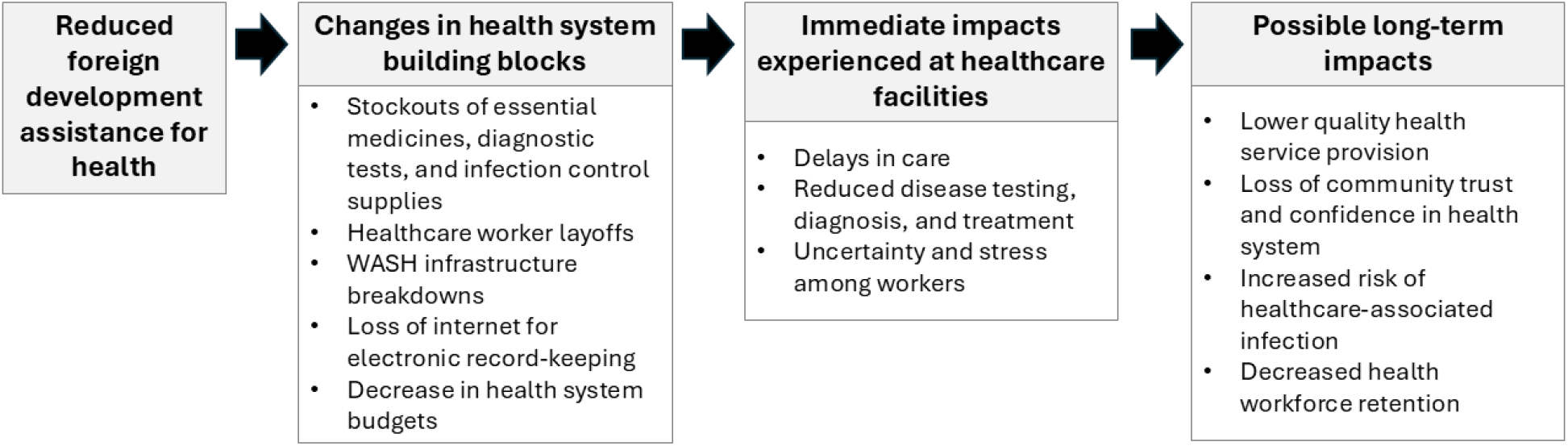
Conceptual framework for changes in health system building blocks, downstream impacts at healthcare facilities, and potential consequences for health arising from reductions in US foreign development assistance.

### Service delivery

Interruptions to health service delivery arose when facilities did not have sufficient materials or staffing levels to provide timely, comprehensive care. Facilities experienced shortages of test kits for HIV, malaria, and tuberculosis, and some participants reported that they had been unable to test patients for these conditions. Laboratory staff also reported shortages of consumable supplies for testing, such as specimen bottles. Timeliness of care was also impacted, with participants reporting that staff layoffs led to longer wait times for patients.

Healthcare facility infrastructure also affected service delivery. Many participants described problems with WASH services at their facilities, such as damage to toilets and ablution blocks, water pump breakdowns, and water pipe issues. These issues were most often reported at rural health posts and health centers. At some facilities, breakdowns in water or sanitation infrastructure were still unaddressed after several weeks or months. In other instances, participants said that district-level health authorities had intervened to make repairs or install new systems. Rather than arising directly from changes in foreign development assistance, infrastructure breakdowns and repair delays may have been attributable to poor operations and maintenance practices that have been observed in low-income healthcare facility settings more generally (30).

> ‘The water system no longer functions… It has affected patient care because sometimes you tell patients to go and wash hands outside instead of doing so within the building.’ – Nurse, rural health center

### Health workforce

Healthcare facilities were affected by worker layoffs due to temporary or ongoing pauses in foreign development assistance. These layoffs particularly impacted staff providing HIV-related services in urban facilities, such as antiretroviral therapy (ART) clinic staff, peer educators, and contact tracers. Participants reported that these departments were now understaffed. In some instances, staff had been laid off and then reinstated, leading to uncertainty around staffing levels and ability to consistently provide services. Community-based health volunteers had received incentive payments from USAID for outreach work related to HIV/AIDS case-finding and treatment. Several participants at rural facilities reported that volunteers were no longer performing their duties after these payments were cut off, reducing community health engagement.

> ‘When the USAID staff were stopped in February, the work got very messy…we depended on their system for so many things.’ – Nurse, urban hospital

### Access to essential medicines and supplies

Many participants reported that issues with essential medicine availability had arisen since the start of 2025. Participants from multiple urban and rural healthcare facilities reported shortages of antimalarial medication, antiretroviral medications, antibiotics, or family planning materials. In some cases, participants reported difficulties in distributing medicines according to the usual schedule or had to allocate smaller quantities to patients due to current or anticipated shortages. One participant reported that their facility lacked funding for transportation to collect medicines from distribution facilities. A May 2025 measure to reduce US funding for medicine procurement in Zambia, separate from USAID program cuts, may have also contributed to changes in medicine availability (31).

> ‘The ARVs [antiretroviral drugs] given to the people above 40 and teenagers have been out of stock. They reduced from giving them for 6 months to 3 months due to anticipated shortage.’ – Nurse, urban hospital

Participants also frequently reported shortages or stock-outs of WASH and infection control supplies, such as surgical gloves, hand soap, and hand sanitizer, at their facilities. Several cleaning staff also reported experiencing shortages of disinfectants and cleaning soaps, which affected their ability to perform job tasks. It was not clear whether infection control supplies had previously been supplied through foreign development assistance programs.

> ‘I am not doing my work like it should be done… Having a shortage of Jik [bleach] makes it difficult for my work.’ – Cleaner, urban hospital

### Health information systems

USAID had supported internet services at some healthcare facilities, and several participants reported that their internet had recently been disconnected. Facilities relied on these internet services for entering electronic medical record data, and participants said that working with records had become more cumbersome due to limited internet access. One participant said that their facility struggled to send health information system reports in a timely manner due to poor internet.

> ‘The Starlink Internet from USAID recently stopped working, it’s been hard to work with records for the facility.’ – Records clerk, rural mini hospital

### Financing and governance

Changes in health financing and governance structures were closely linked. Program cuts made by USAID, and in turn by other multilateral agencies and non-governmental organizations (NGOs) funded by USAID, led to decreases in funding at the healthcare facility level. In addition to layoffs of USAID-supported staff, a few participants said that they had observed decreases in external partnerships or receipt of monetary donations for their programs. Changes in external partnerships were more frequently reported at urban health centers and hospitals compared to rural facilities, potentially due to higher pre-2025 levels of donor participation at large, high-volume facilities.

> ‘From the time [USAID] withdrew, all these NGOs stopped and affected the work.’ – Nurse, urban health center

### Impacts on health and well-being

Patients and healthcare workers experienced downstream impacts from health system disruptions. Several participants expressed concern that patients were not receiving proper treatment due to shortages of essential medicines or testing supplies, potentially endangering health or prolonging illnesses.

> ‘It makes us sad to see patients go back without proper treatment.’ – Laboratory technician, urban health center

Layoffs of USAID-supported staff contributed to higher workloads for the remaining staff, and some participants described feeling fatigued and stressed from attending to high volumes of patients. Funding interruptions and layoffs lowered morale among staff, with some expressing worries about their future job security. Participants also reported increased fear of contracting infections due to the lack of infectious disease testing supplies, WASH supplies, and personal protective equipment at facilities.

> ‘The status of water at the facility is not known. We are not sure if it is safe or contaminated because there’s no equipment for water testing.’ – Laboratory technician, urban hospital

## Discussion

This study assessed changes experienced at healthcare facilities in Zambia in the three to four months following US foreign development assistance program cuts, with healthcare workers reporting impacts across all health system building blocks. Participants described difficulties in providing comprehensive, timely patient care and worries about their own health protection and job security due to the abrupt nature of the cuts. Our assessment revealed that most facilities lacked one or more basic WASH services and experienced stockouts of essential medicines, underscoring the need for improved health infrastructure and strengthened supply chains. Findings provide insight into the consequences of rapid cuts in foreign development assistance for health system strengthening and global health security.

If unaddressed, shortages of diagnostic testing equipment, medicines, and staffing will negatively affect healthcare facilities’ ability to effectively treat patients, ultimately leading to prolonged illness and increased mortality. However, impacts of foreign development assistance program defunding will not be confined to low- and middle-income countries, as weakened health systems pose a threat to global health security (32). Epidemics and pandemics, such as Ebola and COVID-19, amplify and spread in healthcare facilities with little infection control infrastructure (33–35). In our study, healthcare workers reported shortages of diagnostic testing equipment, limited infection control materials, and information system outages, revealing barriers to detection, control, and reporting of diseases with epidemic potential. At a global level, the US withdrawal from the World Health Organization will weaken WHO-coordinated surveillance and response networks, impeding the ability to track and contain epidemics (36). Without coordinated response mechanisms, downstream consequences, within index countries and beyond, are likely to be severe and costly. National health security actors should consider supporting healthcare facility-level infection control infrastructure and global-level epidemic preparedness and response strategies for preventing the next epidemic or pandemic.

Deficiencies in healthcare delivery systems threaten progress toward achieving development targets such as universal health coverage (37). Healthcare facilities’ ability to provide safe, comprehensive care pertains to the United Nations’ 2030 Sustainable Development Goals, including goals for good health and well-being (Goal 3) and universal coverage of clean water and sanitation (Goal 6) (38). Without a consistent revenue source, facilities may struggle to upgrade and maintain medical infrastructure and WASH services, undermining care delivery (30). The government of Zambia has sought to achieve universal health coverage through a strategic plan that includes health workforce expansion, supply chain strengthening, health infrastructure upgrades, and decentralization of services (39,40). However, 42% of Zambia’s per capita health expenditure was donor-funded in 2021; abrupt reductions in external financing will hamper the government’s ability to make and sustain health system investments (41).

Strategies to build health system revenue and resilience can mitigate the impacts of foreign development assistance program defunding. Zambia’s National Health Insurance Management Authority has sought to improve the quality of care through expanded service provision and essential medicine availability, but weak regulation and low public confidence pose barriers to uptake and quality (42–44). Strengthened regulatory oversight of insurance schemes and public outreach could improve revenue generation from insurance, reducing dependence on external donor financing (45). Furthermore, donor financing often targets specific disease areas under a vertical approach, contributing to fragmented financing arrangements (45,46). Integrated healthcare models, where service delivery is coordinated across health sector programs, can reduce costs of care while improving health outcomes (47,48). By identifying areas of overlap across vertical programs and revenue streams, and moving toward greater integration, governments can improve the efficiency of health system operations and overall healthcare spending (49).

Along with revenue generation, governments should also consider resource allocation strategies to mitigate shocks from donor funding withdrawals. The government of Zambia could ring-fence domestic funding for a set of essential health system inputs, ensuring that funds are protected in the case of further external program disruptions (50). This could include support for salaries of essential healthcare staff who were previously donor-funded (e.g., laboratory technicians, data staff, and community health workers) and for maintaining stocks of essential medicines and commodities. Resource allocation strategies should also account for the needs of primary care settings, which provide essential last-mile health services but may contend with weak supply chains and a lack of essential infrastructure (27,51). To support care delivery in these settings, the government could implement facility resilience packages to provide essential WASH services, energy infrastructure, and medical supplies.

Community outreach and engagement can also play a role in maintaining continuity of care and reducing public health impacts of program cancellations. Some of the impacts reported by study participants – such as longer waiting times, reduced availability of medicines, and poor quality of WASH services – may lead to poor community perceptions of healthcare facilities (52,53). Patients who encounter subpar services may feel hesitant to seek care in the future, undermining efforts to increase immunization, attended deliveries, and other facility-based services (54,55). Efforts to build community trust in the health system and maintain service continuity could encourage health-seeking behavior, potentially moderating the health impacts of funding cuts (56,57).

This research is subject to limitations. In face-to-face surveys, participants may have either overstated or minimized the impacts of foreign development assistance defunding due to social desirability bias. To minimize the potential for bias, surveys took place in private locations within healthcare facilities, and researchers received training in impartial interviewing techniques. While interviewers were given a list of probing questions, they were not required to ask all of these questions, potentially introducing bias due to variations in interviewing techniques. We used a purposive sampling approach to select individuals for surveys and only conducted surveys during daytime hours; this approach may be subject to selection bias as we did not include workers who were unavailable or off-duty at the time of data collection. This analysis was intended to assess healthcare worker perspectives on the impacts of foreign development assistance defunding. We did not verify whether the reported changes were directly attributable to USAID program cuts or collect longitudinal data to compare conditions before and after cuts took place. Participants may have reported changes that were unrelated to US-funded programs, or changes that they had heard about but not directly experienced. Due to the timing of data collection, which occurred three to four months after the initial cancellations of US foreign development assistance programs, we were unable to assess long-term changes. To better understand the link between defunding and health system impacts, future research could consult stakeholders involved with USAID programming across multiple levels of the health system in Zambia. Future projects could also employ quasi-experimental approaches to quantify the impacts of funding cuts on health service delivery indicators and health outcomes in affected areas.

## Conclusion

Healthcare facilities in Zambia have experienced interruptions to health service delivery following rapid reductions in US foreign development assistance. Recent studies have modeled the potential health impacts of foreign development assistance defunding or described changes experienced within specific programs (13,17,58), but to our knowledge, ours is the first to document impacts across multiple aspects of a health system at the facility level. Healthcare workers reported changes that impacted their ability to deliver efficient and high-quality care, posing risks to patients’ health and eroding confidence in the healthcare system.

In spite of the disruption caused by funding reductions, this moment presents an opportunity to rethink foreign development assistance priorities and build domestic capacity in recipient countries. To improve the efficiency and sustainability of development initiatives, donors should prioritize capacity-building and aim to integrate programs into the existing health system (59). Country programs should also consider alternatives to conventional development assistance financing models, such as South-South collaboration and multilateral financing frameworks, to work toward shared development priorities. Initiatives to improve health system governance, coordination, and financing can insulate against future disruptions and improve health outcomes in the long term.

## Data Availability

All data produced in the present study are available online in the Carolina Digital Repository.

https://doi.org/10.17615/g2pd-rc04

## Acknowledgements

We acknowledge and thank the healthcare workers who contributed their time and perspectives to this study. We thank Florence Kataya, Kings David Mulenga, and Tenford Siavwemu for their role in data collection. We thank Nikki Benhke for providing feedback on the draft manuscript.

